# Neutralizing and binding antibody kinetics of COVID-19 patients during hospital and convalescent phases

**DOI:** 10.1101/2020.07.18.20156810

**Authors:** Xiang-Yang Yao, Wei Liu, Zhi-Yong Li, Hua-Long Xiong, Ying-Ying Su, Ting-Dong Li, Shi-Yin Zhang, Xue-Jie Zhang, Zhao-Feng Bi, Chen-Xi Deng, Cai-Yu Li, Quan Yuan, Jun Zhang, Tian-Ying Zhang, Zhan-Xiang Wang, Sheng-Xiang Ge, Ning-Shao Xia

## Abstract

Knowledge of the host immune response after natural SARS-CoV-2 infection is essential for informing directions of vaccination and epidemiological control strategies against COVID-19. In this study, thirty-four COVID-19 patients were enrolled with 244 serial blood specimens (38.1% after hospital discharge) collected to explore the chronological evolution of neutralizing (NAb), total (TAb), IgM, IgG and IgA antibody in parallel. IgG titers reached a peak later (approximately 35 days postonset) than those of Nab, Ab, IgM and IgA (20∼25 days postonset). After peaking, IgM levels declined with an estimated average half-life of 10.36 days, which was more rapid than those of IgA (51.25 days) and IgG (177.39 days). Based on these half-life data, we estimate that the median times for IgM, IgA and IgG to become seronegative are 4.59 (IQR 4.12-5.03), 7.78 (IQR 6.71-9.16) and 42.72 (IQR 33.75-47.96) months post disease onset. The relative contribution of IgM to NAb was higher than that of IgG (standardized β regression coefficient: 0.53 vs 0.48), so the rapid decline in NAb may be attributed to the rapid decay of IgM in acute phase. However, the relative contribution of IgG to NAb increased and that of IgM further decreased after 6 weeks postonset. It’s assumed that the decline rate of NAb might slow down to the same level as that of IgG over time. This study suggests that SARS-CoV-2 infection induces robust neutralizing and binding antibody responses in patients and that humoral immunity against SARS-CoV-2 acquired by infection may persist for a relatively long time.

## Introduction

The pandemic caused by SARS-CoV-2 infection is accelerating worldwide, with more than 100 thousand new cases of COVID-19 per day [1]. As of July 12, 2020, more than 12 million cases and 560,000 deaths globally due to the pandemic have been recorded. SARS-CoV-2 infection can be asymptomatic or cause a wide range of symptoms from mildly symptomatic to severe respiratory failure and death [2, 3]. There are several antiviral drug candidates against SARS-CoV-2 in clinical evaluation; however, none have been proven to be effective [4]. In addition, more than 100 vaccines are being developed, with 13 candidates being evaluated clinically [5].

Knowledge of the host immune response after natural SARS-CoV-2 infection is essential for informing the directions of vaccination and epidemiological control strategies against COVID-19. The immune response to SARS-CoV-2 infection among COVID-19 patients has been characterized by several studies [6-11]. Total antibodies (TAb) are detected first, as early as 7 days post exposure or on the day of onset, and the median seroconversion day is reportedly 9 to 11 days postonset [6, 7]. The IgA response appears to occur slightly earlier than that of IgM and IgG, and seroconversion for IgA, IgM and IgG generally occurs after approximately two or three weeks postonset [6-8, 10]. With regard to neutralizing antibodies (NAb), the seropositivity rate can reach up to 100% within 20 days after disease onset [11]. Recent studies indicated SARS-CoV-2 infection may induce transient nAb response, which raised the concern on NAb against re-infection with SARS-CoV-2 and the duration of vaccine protection[12, 13].

However, the majority of relevant studies have been conducted during hospitalization, and the persistence of binding and neutralizing antibodies against SARS-CoV-2 acquired from natural infection remains unknown. Moreover, the direct comparison and the kinetic correlations between NAb and IgG, IgM and IgA are still unclear, since few studies examine all four isotypes in parallel. In this study, we analyzed the chronological evolution of NAb, TAb, IgG, IgM and IgA to gain a greater understanding of the immune response of COVID-19 patients during both hospital and convalescent phases.

## Methods

### Patients

Between January 22 and February 13, 2020, 35 patients with real-time RT-PCR (rRT-PCR)-confirmed SARS-CoV-2 infections were admitted to the Xinglin Branch of the First Affiliated Hospital of Xiamen University (the only designated hospital for COVID-19 in Xiamen), China. Among them, 34 (97.1%) subjects who were willing to donate blood samples were enrolled in this study. All patients were followed semimonthly or monthly after discharge, and blood samples were collected for antibody testing. Personal demographic data, date of illness onset, clinical manifestation and disease classification were obtained from medical records in the hospital.

This study was approved by the Medical Ethical Committee of the First Affiliated Hospital of Xiamen University.

### Pseudovirus-based neutralization assay

The neutralizing activity of serum samples was measured by a previously reported pseudovirus-based neutralization assay, by which the determined neutralizing titers of serum collected from COVID-19 patients correlated well with those according to a live SARS-CoV-2 virus neutralization assay [14]. Briefly, after heat inactivation at 56 □ for 30 min, serum samples were serially diluted twofold, starting from 10-fold dilutions, after which the dilutions were mixed with an equal volume of VSV-SARS-CoV-2-Sdel18 virus (MOI=0.05). All serum samples and viruses were diluted with Dulbecco’s Modified Eagle Medium (Sigma-Aldrich, St Louis, MO, USA) supplemented with 10% fetal bovine serum (GIBCO, Langley, OK, USA). After incubation at 37 °C for 1 hour, 100 μL of the mixture for each dilution was added in duplicate to a cell culture plate previously seeded with BHK21-hACE2 cells, followed by incubation for 12 hours at 37 °C in a humidified atmosphere with 5% CO_2_. Fluorescence images were obtained using an Opera Phenix (PerkinElmer), and GFP-positive infected cells in each well were quantified by the Columbus system (PerkinElmer). The rate of reduction (%) in infection was calculated compared to a control not treated with serum and graphed using GraphPad Prism 8.0 (GraphPad Software Inc., San Diego, California, USA) versus the log of the reciprocal serum dilution. A sigmoidal dose response curve with a variable slope was then generated to determine the 50% neutralization titer (NT_50_), i.e., the serum dilution at which a 50% reduction in infection was observed. In this study, NT_50_ was used as the titer of serum NAb, and samples with NT_50_ ≥ 50 were considered NAb positive.

### Binding antibody testing

TAb, IgG, IgM and IgA against the receptor-binding domain (RBD) of the SARS-CoV-2 spike protein in the sera of COVID-19 patients were tested using commercially available enzyme-linked immunosorbent assay (ELISA) kits, which were manufactured by Beijing Wantai Biological Pharmacy Enterprise Co., Ltd. (Beijing, China). The double-antigen sandwich method and μ-chain capture method were employed to assess TAb and IgM, respectively; indirect immunoassay was used for IgG and IgA. The excellent performance of the kits for detecting TAb and IgM supplied by Wantai has been reported in many studies [6, 7, 15]. All assays were carried out according to the manufacturer’s instructions. Briefly, 100 μL of sample was used for TAb testing, and 10 μL of sample was added to a microwell plate preloaded with 100 μL of sample diluent for the detection of IgM, IgG and IgA. The microplate was incubated at 37 □ for 30 min and washed 5 times with washing buffer; 100 μL of enzyme reagent was then added and incubated at 37 □ for 30 min, followed by another 5 washes. Next, 100 μL of chromogenic substrate was added, and color was developed by incubation at 37 □ for 15 min; the reaction was terminated by the addition of 50 μL of 2 M H_2_SO_4_. Optical density (OD) at 450 nm with 630 nm as a reference was measured, and samples with OD values for a given antibody assay greater than or equal to the cut-off value were determined to be positive for that antibody.

The antibody titer was further determined for samples with positivity for any antibody. Samples were twofold serially diluted with phosphate-buffered saline (pH 7.0) with 20% (v/v) newborn bovine serum, and the twofold dilution set was retested. The greatest dilution of the sample with a positive result was determined, and the antibody titer for this sample was defined as follows: the OD value at this greatest dilution divided by the cut-off value and then multiplied by this dilution factor.

### Statistical analysis

After log transformation, lognormal distribution was used to simulate NAb, Tab, IgG, IgM, and IgA titers against the time postonset. Pearson coefficients of correlation were calculated to relate NAb titers with those of IgM, IgG and IgA at different times after disease onset. Standardized regression coefficients were obtained to examine the independent contributions of IgM, IgG and IgA toward NAb. The temporal change in antibody levels for each patient were calculated by dividing the log-transformed titer difference of two samples collected at adjacent time points by the time interval, except when both samples were negative. For each type of antibody, the mean temporal change in all patients every 5 days against the time postonset was plotted and simulated by four-parameter logistic (4-PL) regression. The value of the intersection of the x-axis and 4-PL curve was deduced as the mean time when a given antibody reached its peak level (the mean of temporal change equaled zero), and the bottom of the 4-PL curve was considered the plateau of the antibody decay. The peak times with peak levels of antibodies were analyzed in participants who donated at least 6 blood samples. All statistical analyses were conducted using SAS 9.4 (SAS Institute, Cary, NC, USA), and graphs were drawn with GraphPad Prism 8.0 (GraphPad Software Inc., San Diego, California, USA).

## Results

### Patient Characteristics

A total of 34 (97.1%, 34/35) patients admitted to the hospital before February 13 were included in this study. As shown in table 1, the median age was 52 (IQR 38, 67), and 58.8% were male. The median duration from onset to hospitalization was 3 days (IQR 2, 6). The majority of patients had fever (76.5%) on admission; cough was the second most common symptom (35.3%). All patents exhibited abnormalities on chest CT. The illness was severe or critical in four patients (11.8%). All patients recovered and were discharged from the hospital before March 3, with a median time from onset to discharge of 23 days (IQR 18, 28). The median follow-up time was 57 days (range 17, 98).

**Table 1.**
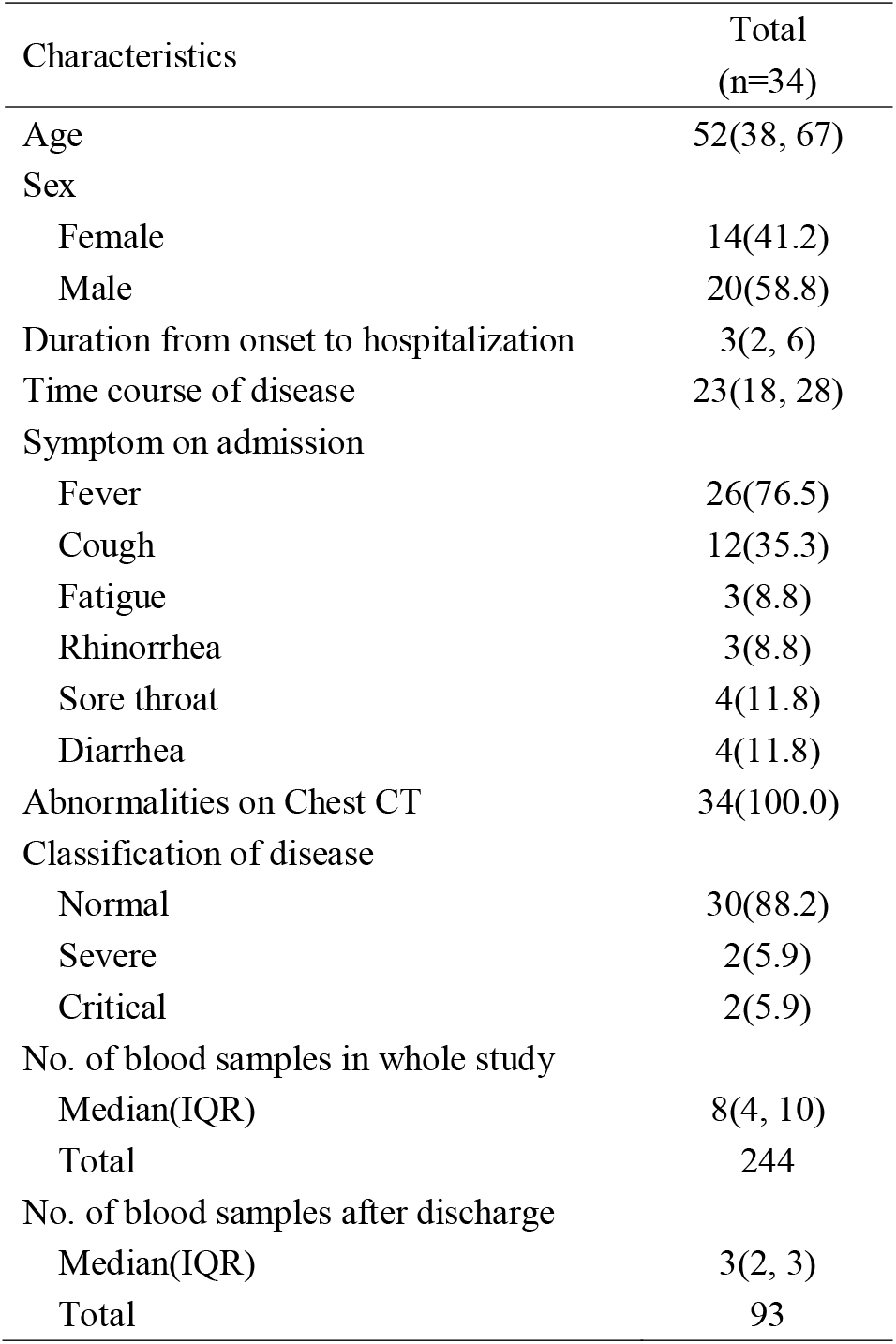
Study Characteristics of participants

### Antibody response against SARS-CoV-2

In total, 244 serial blood specimens were collected from 34 COVID-19 patients (median 8 samples for each patient), with 93 (38.1%) collected after hospital discharge. The responses of NAb, TAb, IgG, IgM and IgA against SARS-CoV-2 are depicted in Figure 1 (the antibody response for each patient is illustrated in Supplementary Figure 1). For each type of antibody, seroconversion occurred in all patients. Before seroconversion, the most recent sample with negative results for NAb, TAb, IgG, IgM and IgA was collected at days 14, 8, 13, 10 and 15 postonset, respectively. Of 77 specimens collected within the first 15 days after onset, 60 (77.92%), 73 (94.81%), 62 (80.52%), 68 (88.31%) and 66 (85.71%) tested positive for NAb, TAb, IgG, IgM and IgA, respectively. No antibody reversion was observed in any patient during the study period, except that IgM in 3 patients became seronegative at days 35, 48 and 54 after onset.

**Figure 1.**
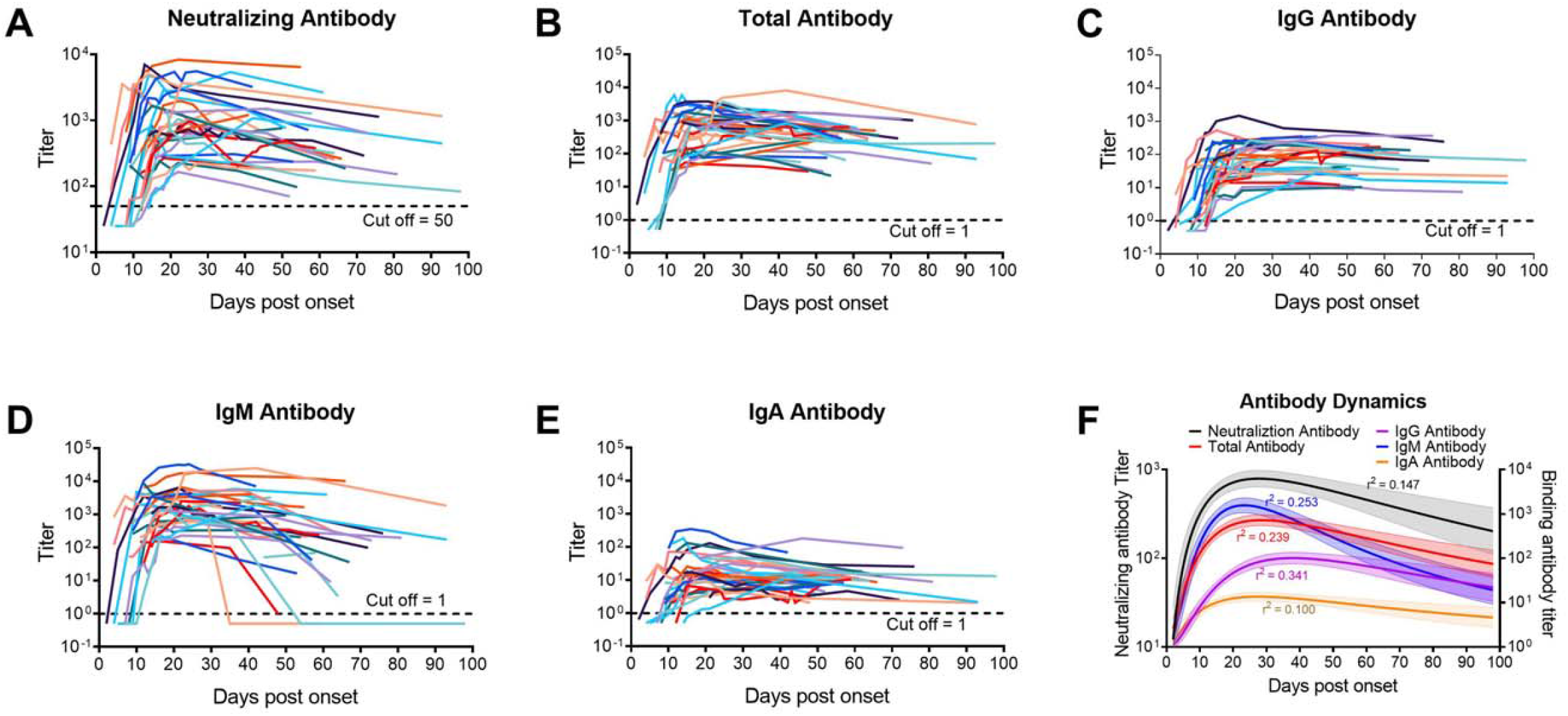
Response of neutralizing, total, IgG, IgM, and IgA antibodies against SARS-CoV-2. (A) Neutralizing antibodies changes of all patients; (B) Total antibodies changes of all patients; (C) IgG antibodies changes of all patients; (D) IgM antibodies changes of all patients; (E) IgA antibodies changes of all patients; (F) Estimated dynamic changes of neutralizing, total, IgG, IgM, and IgA antibodies

### Dynamics of antibodies against SARS-CoV-2

After the onset of illness, antibody tests became positive, and titers increased sharply (Figure 2 and Table 2). The levels of IgM increased most rapidly, followed by TAb, IgG and NAb; IgA titers increased most slowly. Within the first 5 days postonset, the titers of IgM, TAb, IgG, NAb and IgA increased by 2.75 (0.44 log_10_), 2.09 (0.32 log_10_), 1.91 (0.28 log_10_), 1.78 (0.25 log_10_) and 1.48 (0.17 log_10_) times per day, respectively, followed by slower growth. NAb (24.18 days postonset), TAb (23.59), IgM (21.96) and IgA (20.47) reached peak levels at similar times, whereas IgG levels peaked much later (34.49 days postonset). Among peak titers (log_10_), IgM was highest (median 3.36, IQR 2.95-3.74), followed by NAb (median 3.03, IQR 2.86-3.69), TAb (median 2.95, IQR 2.72-3.45) and IgG (median 2.12, IQR 1.66-2.38), with IgA being the lowest (median 1.25, IQR 1.06-1.49) (Table 2).

**Table 2.** Dynamic characteristics of neutralizing and binding antibodies.

|  | Neutralizing<br>Antibody | Total<br>Antibody | IgG<br>Antibody | IgM<br>Antibody | IgA<br>Antibody |
| --- | --- | --- | --- | --- | --- |
| Peak titers ($\log_{10}$, Median, IQR) | | | | | |
|  | 3.03<br>(2.86, 3.69) | 2.95<br>(2.72, 3.45) | 2.12<br>(1.66, 2.38) | 3.36<br>(2.95, 3.74) | 1.25<br>(1.06, 1.49) |
| Time of antibody reaching peak titer (Days postonset) |  |  |  |  |  |
|  | 24.18 | 23.59 | 34.49 | 21.96 | 20.47 |
| Ascending speed (Mean change of $\log_{10}$(titer) per day) | | | | | |
| Day 1-5 | 0.25 | 0.32 | 0.28 | 0.44 | 0.17 |
| Day 6-10 | 0.20 | 0.22 | 0.22 | 0.26 | 0.13 |
| Day 11-15 | 0.08 | 0.09 | 0.14 | 0.10 | 0.05 |
| Day 16-20 | 0.03 | 0.02 | 0.05 | 0.03 | 0.01 |
| Plateau of decay speed (Mean change of $\log_{10}$(titer) per day) | | | | | |
|  | -0.006940 | -0.009895 | -0.001697 | -0.029050 | -0.005874 |
| Half-life of antibody decay (days) |  |  |  |  |  |
|  | 43.38 | 30.42 | 177.39 | 10.36 | 51.25 |
| Time of antibody reversion (Months postonset, Median, IQR) |  |  |  |  |  |
|  | 7.18<br>(6.36, 10.38) | 10.74<br>(9.94, 12.40) | 42.72<br>(33.75, 47.96) | 4.59<br>(4.12, 5.03) | 7.78<br>(6.71, 9.16) |

**Figure 2.**
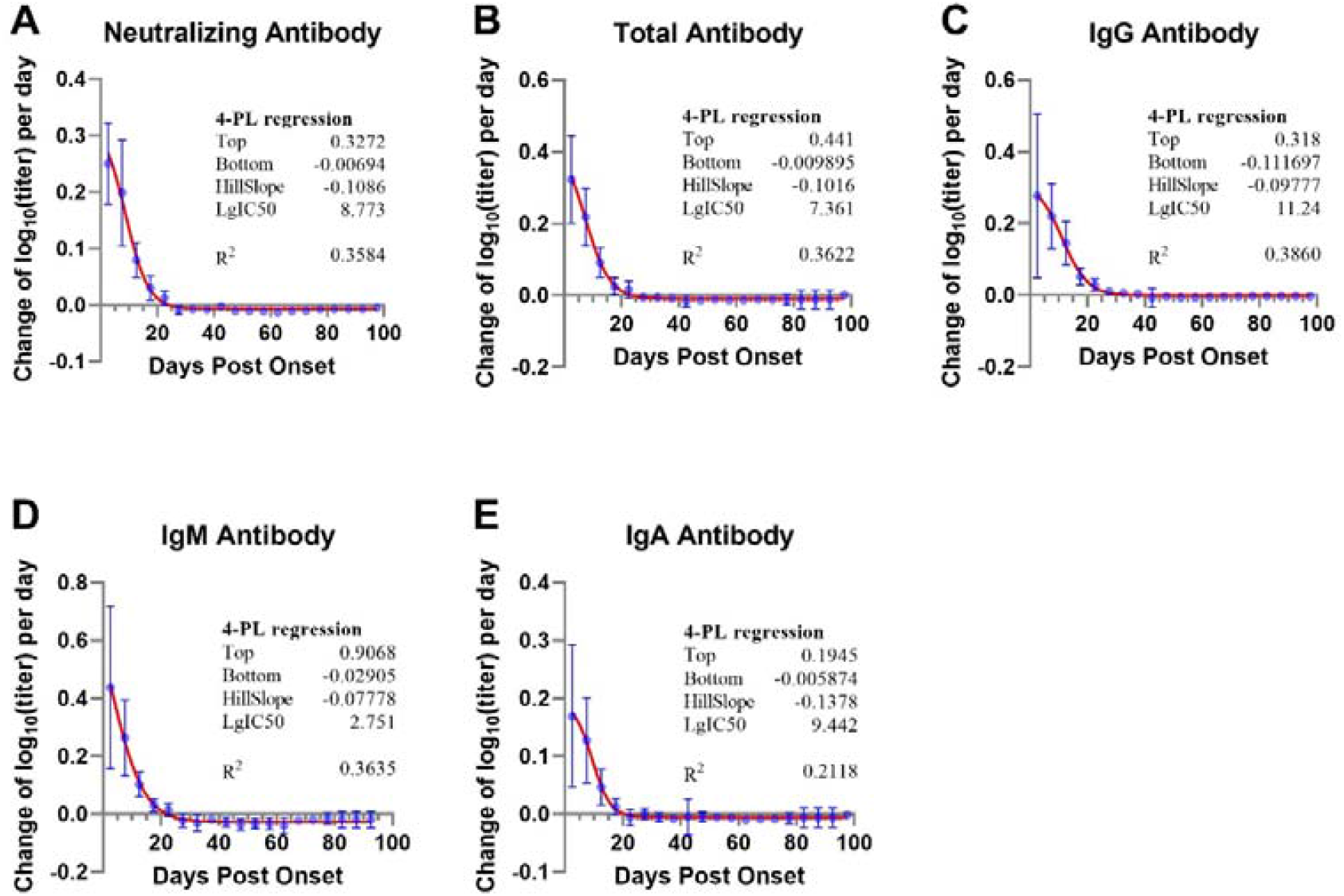
Speed of titer change of neutralizing, total, IgG, IgM, and IgA antibodies against SARS-CoV-2. (A) Neutralizing antibodies; (B) Total antibodies; (C) IgG antibodies; (D) IgM antibodies; (E) IgA antibodies

During the study period, the decay speeds were different for each type of antibody (Figure 2 and Table 2). IgM levels declined the fastest, with a half-life of 10.36 days, followed by IgA (51.25 days) and IgG (177.39 days) levels. The predicted median times for IgM, IgA and IgG antibodies to become seronegative were 4.59 (IQR 4.12-5.03), 7.78 (IQR 6.71-9.16) and 42.72 (IQR 33.75-47.96) months post disease onset. As a mixture of IgG, IgM and IgA, NAb and TAb declined faster than IgG and slower than IgM during the observed period.

### Correlations between neutralizing and binding antibodies

As shown in Tables 3 and 4, significant correlations between NAb and IgG, IgM and IgA were observed, with the highest correlation coefficient between NAb and IgM (r=0.71, 95%CI: 0.64, 0.77, p<0.001), followed by IgG (r=0.59, 95%CI: 0.50, 0.67, p<0.001) and IgA (r=0.37, 95%CI: 0.25, 0.48, p<0.001). The relative contribution of IgM to NAb was higher than that of IgG (standardized β regression coefficient: 0.53 vs 0.48). Because IgM declined more rapidly than did IgG, the relative contribution of IgG to NAb increased after 6 weeks postonset, though the relative contribution of IgM further decreased. The relative contribution of IgM to TAb was higher than that of IgG during the early stage of the disease; however, the relative contribution of IgG was higher than that of IgM six weeks later.

**Table 3.** Correlation between neutralizing, total antibodies and IgG, IgA, IgM antibody

|  | IgG antibody<br>r(95%CI) | IgM antibody<br>r(95%CI) | IgA antibody<br>r(95%CI) | Total antibody<br>r(95%CI) |
| --- | --- | --- | --- | --- |
| Neutralizing antibody |  |  |  |  |
| All serum | 0.59(0.50, 0.67) | 0.71(0.64, 0.77) | 0.37(0.25, 0.48) | 0.72(0.65,0.78) |
| Day 1-14 | 0.80(0.65, 0.88) | 0.79(0.66, 0.88) | 0.49(0.24, 0.68) | 0.75(0.59,0.85) |
| Day 15-28 | 0.71(0.60, 0.80) | 0.86(0.80, 0.90) | 0.39(0.21, 0.55) | 0.80(0.72,0.86) |
| Day 29-42 | 0.38(0.06, 0.63) | 0.68(0.44, 0.82) | 0.12(-0.22, 0.43) | 0.57(0.30,0.76) |
| Day 43- | 0.39(0.10, 0.62) | 0.36(0.04, 0.61) | 0.18(-0.13, 0.46) | 0.47(0.20,0.68) |
| Total antibody |  |  |  |  |
| All serum | 0.68(0.61, 0.75) | 0.74(0.68, 0.80) | 0.45(0.34, 0.55) |  |
| Day 1-14 | 0.80(0.67, 0.88) | 0.90(0.84, 0.94) | 0.64(0.45, 0.77) |  |
| Day 15-28 | 0.70(0.58, 0.79) | 0.77(0.68, 0.84) | 0.38(0.20, 0.54) |  |
| Day 29-42 | 0.46(0.15, 0.68) | 0.73(0.52, 0.85) | -0.01(-0.34, 0.32) |  |
| Day 43- | 0.78(0.62, 0.88) | 0.27(-0.06, 0.54) | 0.36(0.07, 0.60) |  |

**Table 4.** Multiple regression analysis for the association between Neutralizing, total antibody and IgG, IgM and IgA antibodies

| | Standardized $\beta$ regression coefficient | | | $R^2$ |
| --- | --- | --- | --- | --- |
|  | IgG<br>antibody | IgM<br>antibody | IgA<br>antibody |  |
| Neutralizing antibody |  |  |  |  |
| All serum | 0.48 | 0.53 | -0.02 | 0.78 |
| Day 1-14 | 0.59 | 0.44 | -0.05 | 0.88 |
| Day 15-28 | 0.39 | 0.70 | -0.14 | 0.82 |
| Day 29-42 | 0.27 | 0.59 | 0.07 | 0.43 |
| Day 43- | 0.39 | 0.55 | -0.02 | 0.41 |
| Total antibody |  |  |  |  |
| All serum | 0.48 | 0.50 | 0.03 | 0.77 |
| Day 1-14 | 0.25 | 0.73 | 0.002 | 0.89 |
| Day 15-28 | 0.44 | 0.58 | -0.13 | 0.70 |
| Day 29-42 | 0.48 | 0.45 | -0.17 | 0.41 |
| Day 43- | 0.84 | 0.25 | -0.12 | 0.66 |

## Discussion

The present study describes the kinetics of neutralizing and binding antibodies against SARS-CoV-2 during the acute and early convalescent phases of COVID-19 in parallel. Approximately two weeks after illness onset, all types of antibodies had seroconverted, with antibody titers rising sharply. In the early phase, titers of NAb, TAb and IgG on average doubled daily; IgM titers tripled almost daily, and IgA titers increased by an average of 1.5 times per day. The titers of IgG reached a peak later (approximately 35 days postonset) than those of the other antibodies (20-25 days postonset), with IgM decaying with an average half-life of 10.36 days, which was more rapid than IgA (51.25 days) and IgG (177.39 days). Based on these half-life data, we estimate that IgG will last for at least 3.5 years in half of patients, which is similar to the results obtained from patients infected by SARS-CoV-1 [16].

As a respiratory pathogen, SARS-CoV-2 can stimulate a robust response of IgA in infected patients [10, 17]. Yu et al [10] reported that the levels of specific IgA are significantly higher than those of IgM in patients with COVID-19. In our study, we also observed that IgA seroconversion occurred in all patients (Figure 1 and Supplementary Figure 1); however, the response of serum IgA was weaker when compared to IgM and IgG. Among serum IgM, IgG and IgA, the levels of IgA increased most slowly, with the lowest peak titers (Figure 2 and Table 2). In addition, IgA exhibited a weaker correlation with NAb and TAb than did IgG and IgM (Table 3), and almost no relative contribution of IgA to NAb and TAb was observed (Table 4). The above results indicate that the abundance of IgA in the serum of infected individuals is less than that of IgG and IgM. The inconsistency between the results of this study and those of Yu et al may be due to the different assays used.

For many viruses, the specific IgM induced by primary infection play an important role in neutralization [18, 19], though the neutralizing capacity of serum IgM against SARS-CoV-2 is still unknown and needs to be investigated. In this study, multiple regression analysis for the association between NAb versus IgG, IgM and IgA was conducted, and standardized β regression coefficients were employed to evaluate the relative contributions of IgG, IgM and IgA to NAb. The results indicate that the neutralizing activities of sera mainly derive from IgG and IgM. In addition, the relative contributions of IgM were greater than those of IgG after 2 weeks post illness onset during the study period (Table 4). Although the rapid decline in IgM with a higher decay speed than IgG indicates that the relative contribution of IgM to TAb reactivity is less than that of IgG after 6 weeks postonset, IgM still contributed more to neutralizing activity than does IgG in this period. The findings demonstrate that IgM have an important role in serum neutralization activity during the early convalescent phase. Indeed, it is very important to screen convalescent plasma with high neutralization titers for convalescent plasma therapy, which has been reported as a potential effective treatment for COVID-19 [20]. When a high-throughput serological immunoassay, as opposed to a neutralization assay, is required to scale up convalescent plasma production, an assay detecting IgG and IgM simultaneously will be more suitable than detection of IgG or IgM alone. In this study, a higher correlation between TAb and NAb (r=0.47) than between NAb and IgG (r=0.39) was found, as well as between NAb and IgM (r=0.36), in samples collected after 6 weeks postonset (Supplementary Figure 2).

The duration of immunity acquired by infection is an important public health concern and a critical issue for the herd immunity required to prevent and control COVID-19 [21-23]. Recent studies reported that NAb in a certain proportion of recovered COVID-19 patients became negative shortly after disease onset [12, 13]. Short immunity duration may result in annual SARS-CoV-2 outbreaks [21] and also raise widespread concern about the duration of vaccine protection against COVID-19. Based on the estimated decay speeds in this study, we predict that IgG in half of patients will persist for more than 3.5 years (Table 2). Among patients with SARS-CoV-1 and MERS-CoV infection, the decay of antibodies were slowed over time [16, 24, 25]. Therefore, the median IgG duration of 3.5 years is probably an underestimation of the mean decay speed during the early convalescent phase. In the acute infectious period, IgM play an important role in neutralizing activity, so recent studies observed rapid decline in NAb may be attributed to the rapid decay of IgM. It may be not appropriate to use data collected in the early convalescent phase to predict the persistence of NAb. Once IgM become negative, the neutralizing activity of serum will mainly derive from IgG, and the rate of decline of NAb will be the same as that of IgG. As described by Cao et al, the titers of IgG and NAb correlate significantly among patients who recovered from SARS-CoV-1 infection during a 3-year follow-up period (r=0.9) [24]. The data from the present cohort suggest that immunity against SARS-COV-2 in infected individuals will last for several years (more than 3.5 years), which is similar to the duration of the immune response against SARS-CoV-1 infection (at least 3 years) [24].

There are some limitations that should be noted in this study. First, the sample size of 34 patients was small. However, only 35 patients were admitted to our hospital during the study period, and 97.1% of them were enrolled. Second, majority of subjects exhibited mild illness, and only 4 severe/critical cases and no asymptomatic individuals were included. The antibody kinetics of severe/critical and asymptomatic patients likely differ from those observed in this study. Third, due to the convenient collection of blood samples, the sample numbers of each patient varied widely among all patients, and the majority of samples were collected from week 2 to week 4 after illness onset (Supplementary Figure 3). Finally, the follow-up time was short (from 17 to 98 days after onset), and the duration of specific antibodies was estimated only based on the mean rate of decline during the early convalescent phase. The real lasting duration of immunity against SARS-CoV-2 in COVID-19 patients needs to be further investigated in the future with cohort follow-up for several years.

In conclusion, this study suggests that SARS-CoV-2 infection induces robust neutralizing and binding antibody responses in all patients. The mean peak time for IgG was longer than that of IgM and IgA, and the titer of serum IgA was significantly lower than that of IgM and IgG. IgM may play an important role in neutralization during the early convalescent phase, but the contributions of IgG to NAb will increase with the rapid decay of IgM over time. Most importantly, humoral immunity against SARS-CoV-2 acquired by infection is predicted to persist for a relatively long time (at least more than 3.5 years).

## Data Availability

The data used to support the findings of this study are available from the corresponding author upon request.

